# [^18^F]fluorodeprenyl-D2 PET imaging as a novel tool to monitor disease activity in GAD65-Ab Autoimmune Encephalitis

**DOI:** 10.1101/2025.07.23.25331853

**Authors:** Julia S. Dorneich, Jonathan A. Gernert, Lisa Tagnin, Marianthi Zeinaki, Laura Sanzo, Letizia Vogler, Elisabeth Kaufmann, Justina Dargvainiene, Frank Leypoldt, Gérard N. Bischof, Robert Perneczky, Boris-Stephan Rauchmann, Simon Lindner, Günter U. Höglinger, Rudolf A. Werner, Martin Kerschensteiner, Tania Kümpfel, Matthias Brendel, Franziska S. Thaler

## Abstract

**Objectives:** To evaluate [^18^F]fluorodeprenyl-D2 ([^18^F]F-DED) positron-emission-tomography (PET) imaging for monitoring disease activity in autoimmune encephalitis (AIE) associated with glutamic acid decarboxylase 65 (GAD65) antibodies (Abs).

**Methods:** [^18^F]F-DED PET scans were performed in 22 GAD65-AIE patients and 8 controls. [^18^F]F-DED uptake was assessed using dynamic (0-60 min post-injection) and static (30-60 min post-injection) acquisition. [^18^F]F-DED volumes of distribution (VT, 1-tissue-compartment model with carotid input [1TC2k]) and standardized uptake values (SUV) were calculated as a global index of astrogliosis. Furthermore, regional uptake in the cerebellum and mesiotemporal (MT)/ parahippocampal regions was normalized to global cortical/ subcortical uptake (GLM). PET data were correlated with clinical features, MRI findings, and serum biomarkers (neurofilament light chain [sNfL], glial fibrillary acidic protein [sGFAP]).

**Results:** Main clinical phenotypes included limbic encephalitis (LE)/ temporal lobe epilepsy (TLE) (n=14), stiff-person syndrome (SPS) (n=4), cerebellar ataxia (CA) (n=4), with overlapping features in nine individuals. At the time of PET imaging, median age was 55 years, median disease duration was five years, thirteen patients received immunotherapy, and cranial MRI revealed MT swelling or signal changes in five patients (22.7%), and cerebellar atrophy in one patient (4.5%).

Global [^18^F]F-DED uptake was increased in GAD65-AIE patients compared to controls. VT (1TC2k) and SUVr (GLM) analysis revealed significantly higher MT/ parahippocampal [^18^F]F-DED uptake in the entire GAD65-AIE cohort compared to controls. MT/ parahippocampal [^18^F]F-DED uptake was also significantly increased in LE/TLE patients, whereas CA patients showed significantly increased cerebellar [^18^F]F-DED uptake compared to those without CA. No significant correlation was found between [^18^F]F-DED uptake and levels of sNfL in GAD65 target regions, whereas sGFAP levels showed a correlation trend with [¹⁸F]F-DED uptake in the cerebellum and an association with white matter [¹⁸F]F-DED uptake in a voxelwise analysis. [^18^F]F-DED uptake in MT/ parahippocampal regions and cerebellar white matter correlated with clinical severity in LE/TLE and CA patients, respectively.

**Discussion:** [^18^F]F-DED uptake patterns correspond to clinical phenotypes and track disease severity in GAD65-AIE indicating that [^18^F]F-DED PET represents a valuable tool for detection and monitoring disease activity in this patient population.

## Introduction

Autoimmune encephalitis (AIE) associated with autoantibodies against the intracellular enzyme glutamic acid decarboxylase 65 (GAD65) represents one of the most frequent forms of AIE. GAD65-AIE encompasses a spectrum of clinical phenotypes, including limbic encephalitis (LE)/ temporal lobe epilepsy (TLE), cerebellar ataxia (CA), stiff-person-syndrome (SPS), and overlapping manifestations of these entities ^1,2^. The onset of GAD65-AIE can be either subacute or insidious, and treatment responses are often incomplete ^3^. Over the long-term, most patients exhibit persisting clinical deficits, resulting in a chronic disease state with fluctuating disease activity.

Paraclinical tools to monitor disease activity are urgently needed - to help guide decisions and monitor responses to established immunotherapeutic interventions - and to assess the efficacy of novel therapeutic strategies.

Magnetic resonance imaging (MRI) is often of limited utility in diagnosing and monitoring GAD65-AIE, as abnormalities such as mesiotemporal swelling and/ or mesiotemporal T2/fluid attenuated inversion recovery (FLAIR)-hyperintensities are observed in only a minority of patients ^4, 5^. Furthermore, these abnormalities are often missed due to their transient nature and the typically insidious onset of the disease. Consequently, MRI seldomly provides reliable insight into disease activity. In patients with CA, cerebellar atrophy may be observed at later disease stages ^3^, though its presence generally reflects advanced, largely irreversible pathology with limited responsiveness to immunomodulatory therapy.

Positron emission tomography (PET) imaging is increasingly applied in the context of AIE. [^18^F]FDG PET has been evaluated across multiple AIE subtypes – including GAD65-AIE - in a large meta-analysis ^6^, suggesting that [^18^F]FDG PET is a valuable adjunct to MRI due to its high sensitivity to detect AIE-specific metabolic imaging patterns. However, in GAD65-AIE specifically, [^18^F]FDG PET has been report to reveal fewer metabolic abnormalities compared to other AIE subtypes ^7^. In recent years, novel PET tracers with different CNS targets have emerged. For example, translocator protein (TSPO) PET using the radioligand [^18^F]DPA-714, which assesses microglial activation, has been studies in a cohort of 25 AIE patients with different autoantibodies (Abs) (including one patient with GAD65-Abs). This approach showed a positive detection rate of AIE of 72%, compared to 44% using conventional MRI ^8^. Another study using (TSPO) PET in 10 patients with AIE (n=5 with GAD65-Abs) revealed elevated, mostly asymmetric tracer uptake in the mesial temporal lobe of both hemispheres ^9^.

PET imaging with the radiotracer [^18^F]fluorodeprenyl-D2 ([^18^F]F-DED) is a novel technique for evaluating regional monoamine oxidase B (MAO-B) expression. Since MAO-B is predominantly located on the mitochondrial membranes of glial cells and upregulated in reactive astrocytes ^10^, [^18^F]F-DED PET enables in vivo assessment of reactive astrogliosis ^11^, a feature commonly observed in neuroinflammatory and neurodegenerative conditions. Astrocytes as the major cellular source of an increased MAO-B-PET signal was previously confirmed ^12^. A transgenic mouse model of Alzheimeŕs disease demonstrated that [^18^F]F-DED signal elevation preceded changes detected by TSPO PET and β-amyloid PET ^11^. Additionally, strong [^18^F]F-DED binding was observed in a patient with autoimmune encephalitis, suggesting that [^18^F]F-DED PET may be a promising tool for evaluating the regional extent of inflammation in AIE ^11^.

In line with CARE guidelines, we therefore conducted a PET imaging study using [^18^F]F-DED to cross-sectionally compare tracer uptake in patients with GAD65-AIE versus a control cohort. We further correlated [^18^F]F-DED uptake with clinical phenotype, disease severity, as well as with biomarkers indicative of neuroaxonal damage and astroglial activation.

## Methods

### Study design and Cohorts

This observational, monocentric study included n=22 subjects with GAD65-AIE, presenting with limbic encephalitis (LE)/ temporal lobe epilepsy (TLE), cerebellar ataxia (CA), or stiff person syndrome (SPS). Patients were diagnosed according to proposed diagnostic criteria ^3^ and followed up by neurologists specialized in AIE treatment at the Institute of Clinical Neuroimmunology, LMU Hospital. Clinical data, including the Montreal Cognitive Assessment (MoCa), the modified Rankin Score (mRS), and the Scale for the Assessment and Rating of Ataxie (SARA), were collected at the time of PET imaging (**Fig. 1 A**). To assess clinical severity, composite scores were used based on a shortened, GAD65-AIE-tailored version of the clinical assessment scale in autoimmune encephalitis (CASE) score ^13^, as follows: LE:

- *Mild:* No seizures or seizures stopped ≥ 3 months before PET imaging; no cognitive impairment or cognitive impairment only subjectively reported and not detectable by testing.
- *Moderate*: Ongoing seizures < 3 months before PET imaging or cognitive impairment detectable in testing.
- *Severe*: Ongoing seizures < 3 months before PET imaging and cognitive impairment detectable in testing.

**Figure 1:**
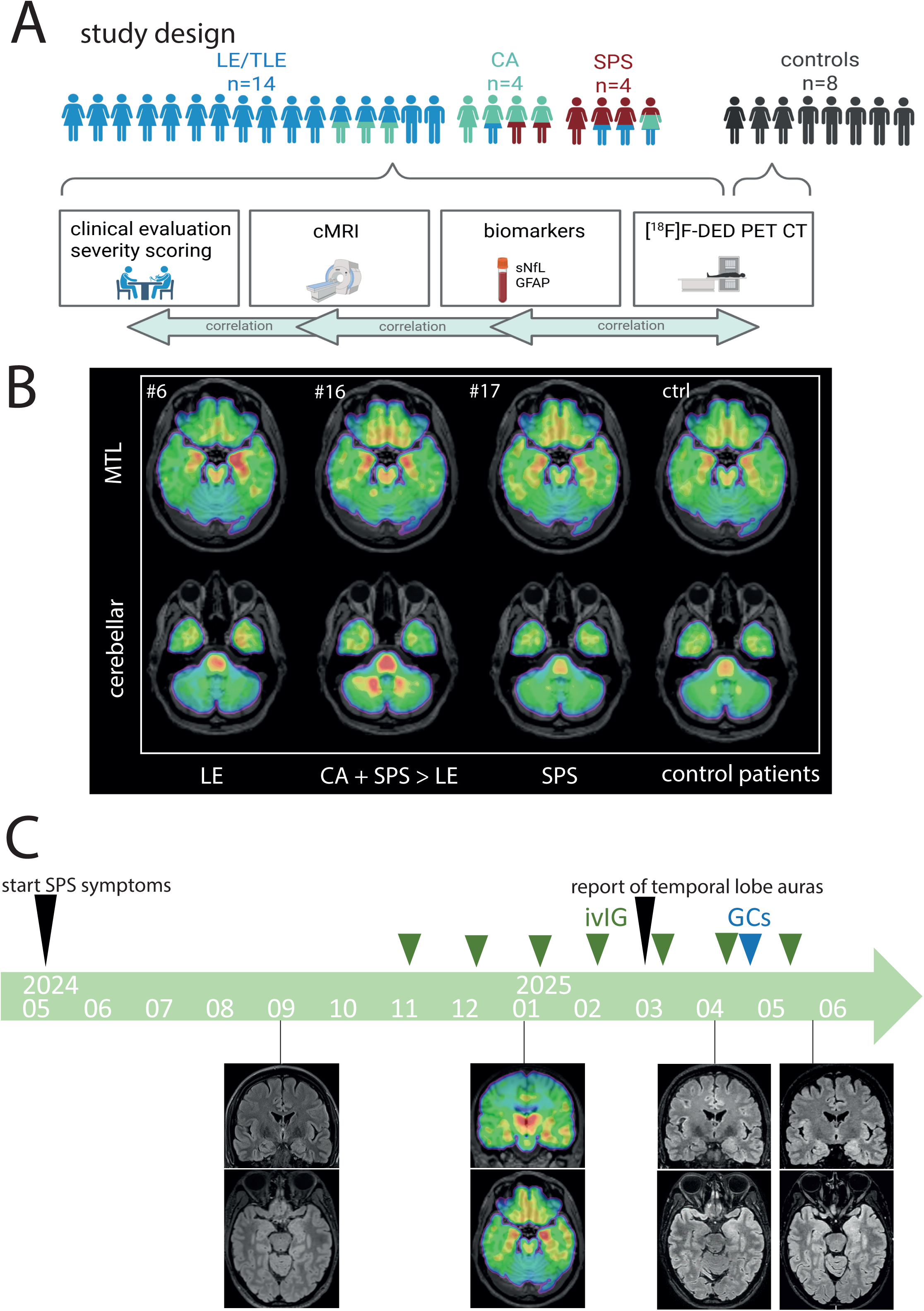
Study overview and qualitative assessment of regional [18F]F-DED uptake. (**A**) Overview of study participants with different clinical phenotypes depicted in distinct colors: limbic encephalitis (LE)/ temporal lobe epilepsy (TLE): blue; cerebellar ataxia (CA): green; stiff person syndrome (SPS): red, including overlapping clinical features with an overview of the analyzed parameter; generated in Biorender. (**B**) [^18^F]F-DED uptake as visualized by SUVr with global mean scaling, is shown for three representative patients with GAD65-AIE: patient #6 with LE/TLE, showing mesiotemporal [^18^F]F-DED uptake; patient #16 with overlapping clinical features of CA, SPS, and LE, showing both mesiotemporal and cerebellar [^18^F]F-DED uptake, and patient #17 with isolated SPS showing no increased [^18^F]F-DED uptake). Additionally the average [^18^F]F-DED uptake of eight controls is shown on the right. (**C**) The case of a patient (#21) initially presenting with SPS symptoms and normal cMRI findings is illustrated. [^18^F]F-DED PET imaging revealed bilateral (left-predominant) mesiotemoporal and parahippocampal [^18^F]F-DED uptake, which preceded the clinical onset of temporal lobe epileptic activity, as well as swelling and FLAIR hyperintensity of amygdalae and parahippocampal regions on cMRI.

CA:

- *Mild:* Cerebellar signs detectable on clinical examination but not perceived by patient.
- *Moderate*: Clinically affected, no walking aid required.
- *Severe*: Walking aid required.

SPS:

- *Mild:* Only signs detectable on examination, such as brisk reflexes or increase startle reaction, without daily activity interference.
- *Moderate:* Clinically affected; no walking aid required.
- *Severe*: Walking aid required, at least outside of buildings.

[^18^F]F-DED PET was performed for diagnostic purposes in all GAD65-AIE patients between 06/2022 and 02/2025. The control cohort included 8 individuals (3 females, 5 males; median age at imaging: 59.5 years). Diagnoses in the control cohort included subjective cognitive dysfunction (n=5), transient neuropsychiatric symptoms (n=1), oligodendroglioma WHO °II (n=1), and unclear transient episodes with decreased consciousness, of unclear etiology (n=1). All subjects provided written informed consent. The study was conducted in accordance with the principles of the Declaration of Helsinki and was approved by the ethics committee of the Ludwig Maximilian University Munich (IRB numbers: 21-0721, 22-0997) and the German radiation protection authorities (Bundesamt für Strahlenschutz (BfS); ZD 3-22464/2023-042-G).

### Antibody testing

GAD65-antibody testing in serum and cerebrospinal fluid (CSF) was performed by indirect immunofluorescence (IIFT) using transfected HEK cells (Institute of Clinical Chemistry, EUROIMMUN Medizinische Labordiagnostika AG, Lübeck, Germany). Intrathecal GAD65-Ab production was defined by a specific antibody index of > 4.

### sNfL and sGFAP measurements

Serum sampling was performed within 6 months of PET imaging (except in patient #8 the interval was 28 months). Serum neurofilament light chain (sNfL) and serum glial fibrillary acidic protein (sGFAP) levels were quantified using the Simoa technology (Neurology 2-Plex B assay; sNfL, sGFAP) on the HX-analyzer as single measurements at the Institute of Clinical Chemistry, Universitätsklinikum Schleswig-Holstein, Campus Kiel.

### cMRI evaluation

Clinical routine cMRI scans of the GAD65-AIE cohort performed within 6 months of PET imaging, as well as scans from the past disease history, were evaluated by a neurologist experienced in AIE-related imaging. MRI scans were acquired using different scanners but allowed standardized qualitative evaluation of: I) Atrophy or swelling of the mesiotemporal (MT) lobe or cerebellum. II) Signal alterations in T2/ FLAIR and T1 sequences of the MT lobe or cerebellum.

### [^18^F]F-DED PET imaging

Dynamic [^18^F]F-DED PET scans (0-60 minutes post-injection) (mean: 166.9 MBq, SD: 23.0 MBq, range: 108.9 MBq) were performed at the Department of Nuclear Medicine, LMU University Hospital, as described previously ^11^. Spatial normalization via structural MRI scans was achieved using PMOD. Volumes of distribution (VT) were determined using a one-tissue-compartment model (1TC2k) with a carotid image-derived input function. For simplified quantification, standardized uptake values (SUV) were extracted from a late time window (30-60 minutes post-injection). In three patients with GAD65 AIE only the late static scan was performed due to more severe disability and the need for reduced scanning time. Regions of interest – including the MT lobe, parahippocampal region, and cerebellar white matter ^14^ – were manually defined in PMOD, whereas a predefined composite of cortical and subcortical regions served as a reference for global astrogliosis assessment. An exploratory PET-to-serum-biomarker correlation analysis was performed using a voxel-wise regression model (sNfL and sGFAP as predictors) in statistical parametric mapping (SPM) implemented in MATLAB (v2011-R2016).

### Statistical Analysis

GraphPadPrism (Version 10) was used for statistical analysis, and Biorender and Adobe Illustrator for figure creation. P-values of < 0.05 were considered statistically significant. Normal distribution was tested using the Shapiro-Wilk test. For multi-comparison testing, groups were compared by Kruskal-Wallis-test followed by Dunńs multiple comparisońs test or ANOVA followed by Tukey’s multiple comparisons test. Comparison of two groups was performed by unpaired T-test. Correlations were assessed using linear-regression. Positive detection rates of MT abnormalities in cMRI and [^18^F]F-DED PET imaging were compared by Fisheŕs exact test.

## Data Availability

Data are available to qualified researchers upon reasonable request.

## Results

### Study Cohort

A total of 22 patients with GAD65-AIE were included in the study (n=14 with LE/TLE, n=4 with SPS, n= 4 with CA, with overlapping clinical phenotypes in nine individuals) (**Fig. 1A**). As expected in GAD65-AIE, there was a strong female predominance (n=20). The median age at PET imaging was 55 years and the median disease duration was five years. At the time of PET imaging, nine patients were untreated, while 13 were receiving immunomodulatory or immunosuppressive treatment, as specified in **Table 1**. At time of diagnosis all patients revealed high-titer GAD65-Ab titers in serum (median: 1:12800) and CSF (median: 1:200) with evidence of intrathecal GAD65-Ab synthesis (**Table 1**).

**Table 1:** Overview of GAD65-AIE cohort at time of [^18^F]fluorodeprenyl-D2 PET imaging.

| Patient ID | dominant phenotype | overlapping phenotype | sex | age (range) * | disease duration (y) * | treatment <sup>§</sup> | GAD65-Ab titer (serum), 1:X * | GAD65-Ab titer (CSF), 1:X * | mRs | SARA | MOC A | LE severity " | CA severity " | SPS severity " | cMRI abnormalities<sup>§</sup> (MT or cerebellar) | Past cMRI abnormalities (MT or cerebellar) |
| --- | --- | --- | --- | --- | --- | --- | --- | --- | --- | --- | --- | --- | --- | --- | --- | --- |
| #1 | LE | / | f | 51-55 | 12 | MMF | 1600 | 50 | 2 | 0 | 26 | 3 | 0 | 0 | MT swelling: UL | MT swelling: UL |
| #2 | LE | / | f | 41-45 | 1 | none | 25600 | 200 | 1 | 0 | nd | 1 | 0 | 0 | none | none |
| #3 | LE | / | f | 31-35 | 5 | none | 6400 | 50 | 2 | 0 | 30 | 3 | 0 | 0 | none | MT swelling: BL |
| #4 | SPS | LE | f | 56-60 | 13 | ivIG, MMF | 2560 | nd | 1 | 1 | 30 | 1 | 0 | 2 | nd & | none |
| #5 | LE | / | f | 26-30 | 8 | none | 1600 | 100 | 1 | 0 | 30 | 2 | 0 | 0 | none | none |
| #6 | LE | CA | f | 56-60 | 5 | none | 3200 | 100 | 2 | 1 | 27 | 2 | 1 | 0 | none | MT swelling: BL |
| #7 | CA | SPS | f | 61-65 | 2 | rituximab | 10 | 10 | 3 | 16 | nd | 0 | 3 | 1 | none | none |
| #8 | LE | / | f | 26-30 | 8 | none | 400 | 50 | 2 | 1 | 29 | 3 | 0 | 0 | nd | MT swelling: UL |
| #9 | LE | / | f | 46-50 | 9 | GCs, PLEX | 3200 | 100 | 1 | 0 | 28 | 3 | 0 | 0 | none | MT swelling: UL |
| #10 | LE | CA | f | 31-35 | 9 | rituximab | 800 | 50 | 2 | nd | 26 | 3 | 2 | 0 | MT swelling: BL | MT swelling: BL |
| #11 | LE | / | m | 41-45 | 4 | none | 400 | 100 | 2 | nd | nd | 2 | 0 | 0 | none | none |
| #12 | LE | / | f | 26-30 | 3 | GCs, PLEX | 12800 | 100 | 1 | 0 | 29 | 2 | 0 | 0 | MT swelling: UL | MT swelling: BL |
| #13 | CA | SPS | f | 71-75 | 12 | PLEX, MTX | 6400 | nd | 3 | 12.5 | 27 | 0 | 3 | 1 | none | none |
| #14 | CA | LE | f | 81-85 | 20 | none | 3200 | nd | 2 | 8.5 | 22 | 2 | 2 | 0 | nd | none |
| #15 | LE | / | m | 61-65 | 3 | GCs | 25600 | nd | 1 | 1 | 25 | 2 | 0 | 0 | none | MT swelling: UL |
| #16 | SPS | CA, LE | f | 56-60 | 9 | ivIG | 25600 | 1600 | 2 | 3.5 | 28 | 1 | 2 | 3 | cerebellar atrophy | cerebellar atrophy |
| #17 | SPS | / | f | 56-60 | 1 | ivIG, ustekinumab<sup>#</sup> | 1600 | nd | 3 | nd | 27 | 0 | 0 | 3 | none | none |
| #18 | LE | CA | f | 81-85 | 1 | none | 12800 | 100 | 2 | 7 | 9 | 2 | 2 | 0 | none | none |
| #19 | CA | / | f | 56-60 | 3 | Aza | 6400 | 100 | 1 | 3.5 | nd | 0 | 2 | 0 | none | none |
| #20 | LE | / | f | 61-65 | 0 | none | 25600 | 100 | 0 | 0 | 30 | 1 | 0 | 0 | none | MT swelling: UL |
| #21 | SPS | LE | f | 36-40 | 0 | ivIG | 25600 | nd | 1 | 0 | 30 | 2 | 0 | 2 | MT swelling: UL | none |
| #22 | LE | / | f | 11-15 | 1 | GCs | 6400 | 100 | 0 | 0 | nd | 1 | 0 | 0 | MT swelling: UL | MT swelling: UL |
| median |  |  |  | 55 | 5 |  | 6400 | 100 | 2.0 | 1.0 | 28 | 1 | 0 | 0 |  |  |
| IQR |  |  |  | 25.3 | 6.8 |  | 20800 | 50 | 1.0 | 3.5 | 4 | 1 | 2 | 0 |  |  |

\* at time of PET imaging; <sup>§</sup> within 3 months prior to PET imaging; <sup>#</sup> due to comorbidities; <sup>§</sup> within 6 months of PET imaging; & due to cardiac pacemaker; " mild: 1, moderate: 2, severe: 3; Aza = azathioprine; BL = bilateral; CA = cerebellar ataxia; cMRI = cranial magnet resonance imaging; f = female; GCs = glucocorticosteroids; ivIG = monthly intravenous immunoglobulins; IQR = interquartile range; MMF = mycophenolate-mofetile; MOCA = Montreal cognitive assessment scale; mRs = modified Rankin scale; MT = mesiotemporal; MTX = methotrexate; nd = no data; LE = limbic encephalitis; m= male; PLEX = plasma exchange; SARA = scale for assessment and rating of ataxia; SPS = stiff person syndrome; UL = unilateral.

Clinical severity was assessed using several standardized scales. The median modified Rankin Scale (mRS) score was 2.0, indicating moderate disability. The median Scale for the Assessment and Rating of Ataxia (SARA) score was 1.0, and the median Montreal Cognitive Assessment (MoCA) score was 28 out of 30, suggesting minimal cognitive impairment in most patients. Severity ratings based on composite clinical scores (see Methods) were as follows:

- LE as main manifestation: *Mild*: n=3, *Moderate*: n=6, *Severe*: n=5.
- LE as overlapping manifestation: *Mild*: n=2, *Moderate:* n=2.
- CA as main manifestation: *Moderate*: n=2, *Severe*: n=2.
- CA as overlapping manifestation: *Mild:* n=1, *Moderate:* n=3.
- SPS as main manifestation: *Moderate*: n=2, *Severe*: n=2.
- SPS as overlapping manifestation: *Mild*: n=2 (**Table 1**).

At the time of PET imaging, MT lobe abnormalities were observed in five of 22 patients (swelling and/ or FLAIR hyperintensity; no MT atrophy was observed), and cerebellar atrophy in one patient. Over the past course of disease, 10 of 22 patients had shown MT lobe abnormalities on cMRI. Further imaging and clinical data are detailed in **Table 1**.

### Qualitative Assessment of [^18^F]F-DED PET Imaging in GAD65-AIE

We first assessed whether [^18^F]F-DED uptake is increased in GAD65-AIE patients compared to controls, and whether regional uptake patterns correspond with clinical phenotypes. Visual analysis revealed distinct uptake patterns: in patients with LE/TLE we observed uptake in the MT/ parahippocampal structures; in patients with CA we observed uptake in the cerebellar white matter; in patients with overlapping LE/TLE and CA we observed focal uptake in both areas; in patients with isolated SPS no increased [^18^F]F-DED uptake was observed (**Fig. 1B**).

One patient (#21) that presented with SPS symptoms in May 2024 and showed no MT abnormalities on cMRI in September 2024, subsequently demonstrated pronounced MT uptake in [^18^F]F-DED PET imaging in January 2025. By March 2025, the patient reported déjà-entendu and subsequently déjà-vu sensations consistent with temporal lobe auras. A follow-up cMRI in April 2005 showed left-sided amygdala and hippocampal swelling and FLAIR-hyperintensities, followed by bilateral MT signal changes in May 2025. Thus, [^18^F]F-DED PET abnormalities preceded both clinical and cMRI evidence of LE in this case (**Fig. 1C**).

### Quantitative Assessment of [^18^F]F-DED PET by Region and Phenotype

We next quantified [^18^F]F-DED uptake in patients with GAD65-AIE and controls using dynamic VT values (1TC2k model with carotid-derived input function). Global VT signal was significantly higher in GAD65-AIE patients than in controls (*p*=0.0228) (**Fig 2A**). Furthermore, GAD65-AIE patients showed significantly increased [^18^F]F-DED uptake both in the cerebral cortex (*p*=0.035) as well as in the MT/ parahippocampal region (*p*=0.019) (**Fig. 2B**). When comparing different clinical phenotypes (dominant clinical phenotype), a significant increase of [^18^F]F-DED uptake in the MT/ parahippocampal region was observed in patients with LE/TLE compared to controls (*p*=0.0166), but not in patients with CA or SPS (**Fig. 2C**). To assess regional uptake differences independent of global uptake levels, we furthermore used SUV values normalized to global cortical and subcortical uptake (SUVr GLM) (**eFig 1A**). SUVr GLM values correlated with VTr GLM values (*p*<0.0001) (**eFig 1B**), suggesting that blood flow was not a confounder of static late-phase imaging. SUVr GLM analysis showed increased MT/parahippocampal uptake in GAD65-AIE patients compared to controls (*p*=0.004). No significant differences were observed in cerebellar uptake (**Fig. 2D**). However, when analyzing all patients with CA (including overlapping phenotypes), cerebellar white matter uptake was significantly increased compared to non-CA patients (*p*=0.0387) (**Fig. 2E**).

**Figure 2:**
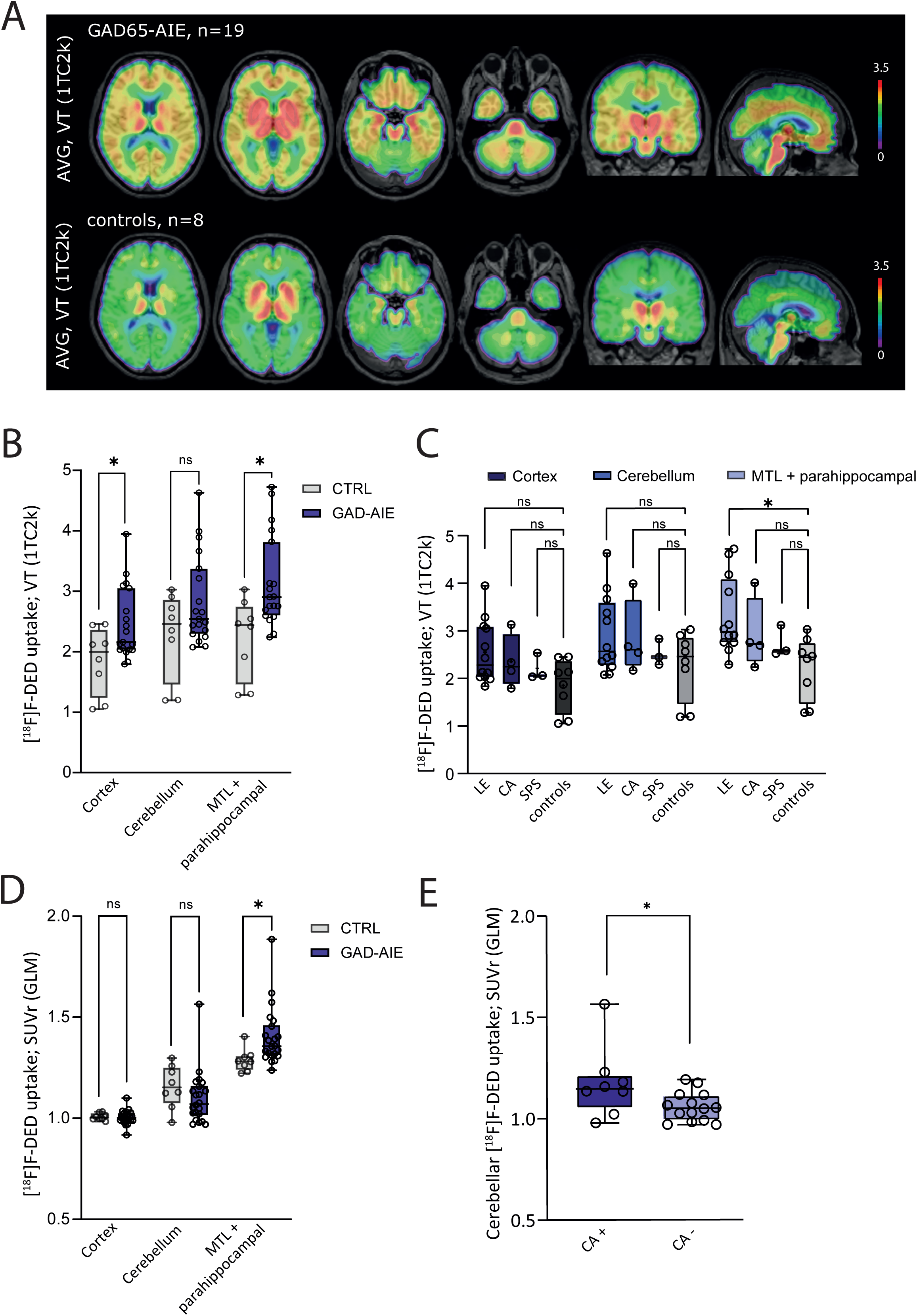
Quantitative assessment of [18F]F-DED uptake. (**A**) Global [^18^F]F-DED uptake, quantified by volumes of distribution (VT, 1TC2k model), is shown as an average for GAD65-AIE patients (n=19) and controls (n=8). VT values could not be calculated for 3 GAD65-AIE patients due to the absence of dynamic imaging. (**B**) [^18^F]F-DED uptake was quantified by VT and compared between GAD65-AIE patients and controls in the cortex, mesiotemporal (MT)/parahippocampal region, and cerebellar white matter. Unpaired T-tests with Welch-correction were used to determine differences. (**C**) [^18^F]F-DED uptake (VT), in the cortex, MT/parahippocampal region, and cerebellar white matter was compared across different clinical phenotypes (dominant clinical phenotype). Normality testing was assessed using the Shapiro-Wilk test. As normality testing was passed, one-way ANOVA followed by Tukeýs multiple comparison test was applied. (**D**) [^18^F]F-DED uptake (standard uptake values [SUVr] with global mean scaling) in the cortex, MT/parahippocampal region, and cerebellar white matter was compared across different clinical phenotypes (dominant clinical phenotype). Normality testing was assessed using the Shapiro-Wilk test. The Kruskal-Wallis test followed by Dunńs multiple comparisońs test was used for the MT/parahippocampal region and cerebellum (due to non-normal distribution), while a one-way ANOVA followed by Tukeýs multiple comparison test was applied for the cortex data (normally distributed). (**E**) Cerebellar [^18^F]F-DED uptake (SUVr with global mean scaling) was compared between patients with cerebellar ataxia (CA) (as either a dominant and overlapping phenotype) and those without CA. As a normal distribution was detected, an unpaired T-test was applied.

Comparison of regional [^18^F]F-DED uptake between treated and untreated patients showed no significant differences (**eFig. 2A**). No correlation was found between uptake in MT/ parahippocampal or cerebellar white matter regions and disease duration in LE/TLE or CA patients, respectively (**eFig. 2 B, C**).

### Correlation of [^18^F]F-DED Uptake with Biomarkers

Next, we evaluated the correlations between [^18^F]F-DED uptake and the serum levels of biomarkers potentially reflecting disease activity in patients with GAD65-AIE: serum neurofilament light chain (sNfL) as a marker for neuroaxonal damage, and serum glial fibrillary acidic protein (sGFAP) as a marker for astroglial activation. No significant correlations were observed between sNfL levels and regional [^18^F]F-DED uptake in cortex, MT/parahippocampal structures, or cerebellum (**Fig. 3A**) nor when applying an exploratory voxel-wise analysis (**Fig. 3C, D**). For sGFAP, a correlation trend of cerebellar tracer uptake and sGFAP levels was observed, while no significant correlation with cortical and MT/parahippocampal uptake was observed (**Fig. 3B**). Furthermore, an exploratory voxel-wise analysis revealed an association of [^18^F]F-DED correlations with sGFAP levels with predominance in the subcortical white matter, basal ganglia, cerebellar white matter, and pons (**Fig. 3C, D**).

**Figure 3:**
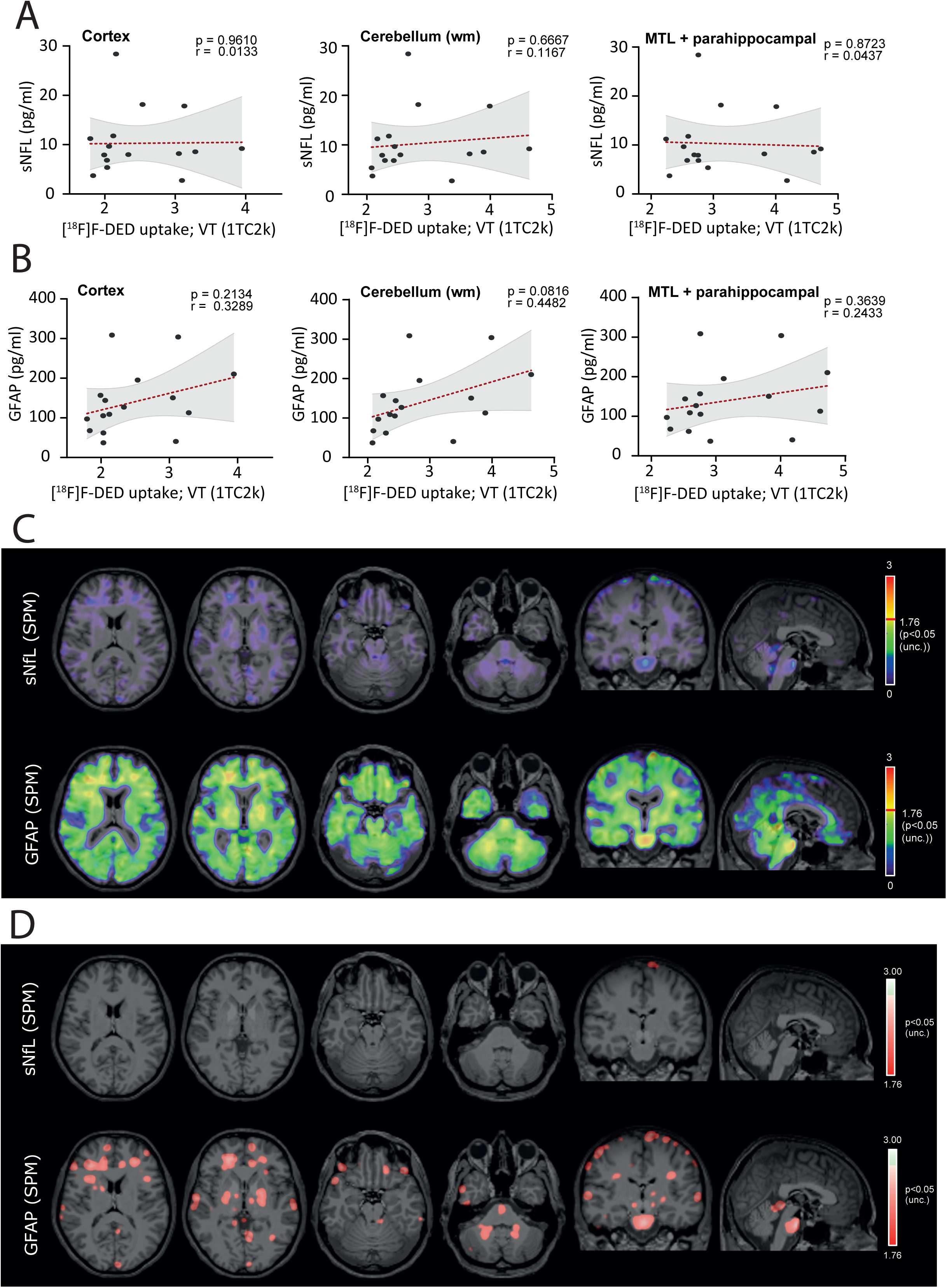
Correlation of [18F]F-DED uptake with serum biomarkers. Correlation of [^18^F]F-DED uptake (volumes of distribution [VT], 1TC2k model) in the cortex, mesiotemporal (MT)/parahippocampal region, and cerebellar white matter with levels of serum neurofilament light chain (sNfL) (**A**), and serum glial fibrillary acidic protein (sGFAP) (**B**) was assessed using linear-regression. (**C, D**) Statistical parametric mapping (SPM) of VT-values was performed to identify voxel-wise positive associations with sNfL and sGFAP levels; the p-value threshold for t-scores that corresponds to α = 0.05 is indicated by the red stroke (**C**). In (**D**) Only voxels that meet the statistically significant T-value threshold as defined in (**C**) are shown in color.

### Correlation of [^18^F]F-DED uptake with Clinical Severity and cMRI findings

To evaluate concordance between clinical severity and astroglia PET imaging, we generated a heatmap of clinical and PET phenotypes and severity grades (**Fig. 4A**). Except for two patients (#7 and #19), PET signal matched the clinical presentation. Patient #7 had cerebellar ataxia without cerebellar tracer uptake. This patient – additional to GAD65-AIE - had previously been diagnosed with CLL and started treatment with rituximab 1,5 years prior to PET imaging, achieving clinical stabilization, GAD65-Ab clearance, and no disease progression - suggesting complete treatment response. Patient #19 had clinical CA with cerebellar tracer uptake, but also MT/ parahippocampal uptake. In this patient due to a language barrier, signs of LE may have been unnoticed. In all other cases, PET uptake corresponded to clinical phenotype. Notably, [^18^F]F-DED PET detected MT abnormalities in 83.3% (15/18) of GAD65-LE/TLE patients, compared to 27.8% (5/18) via cMRI (*p*= 0.002). The three remaining GAD65-LE/TLE patients without MT/ parahippocampal PET uptake reported only subtle LE symptoms (clinical severity grade: 1)

**Figure 4:**
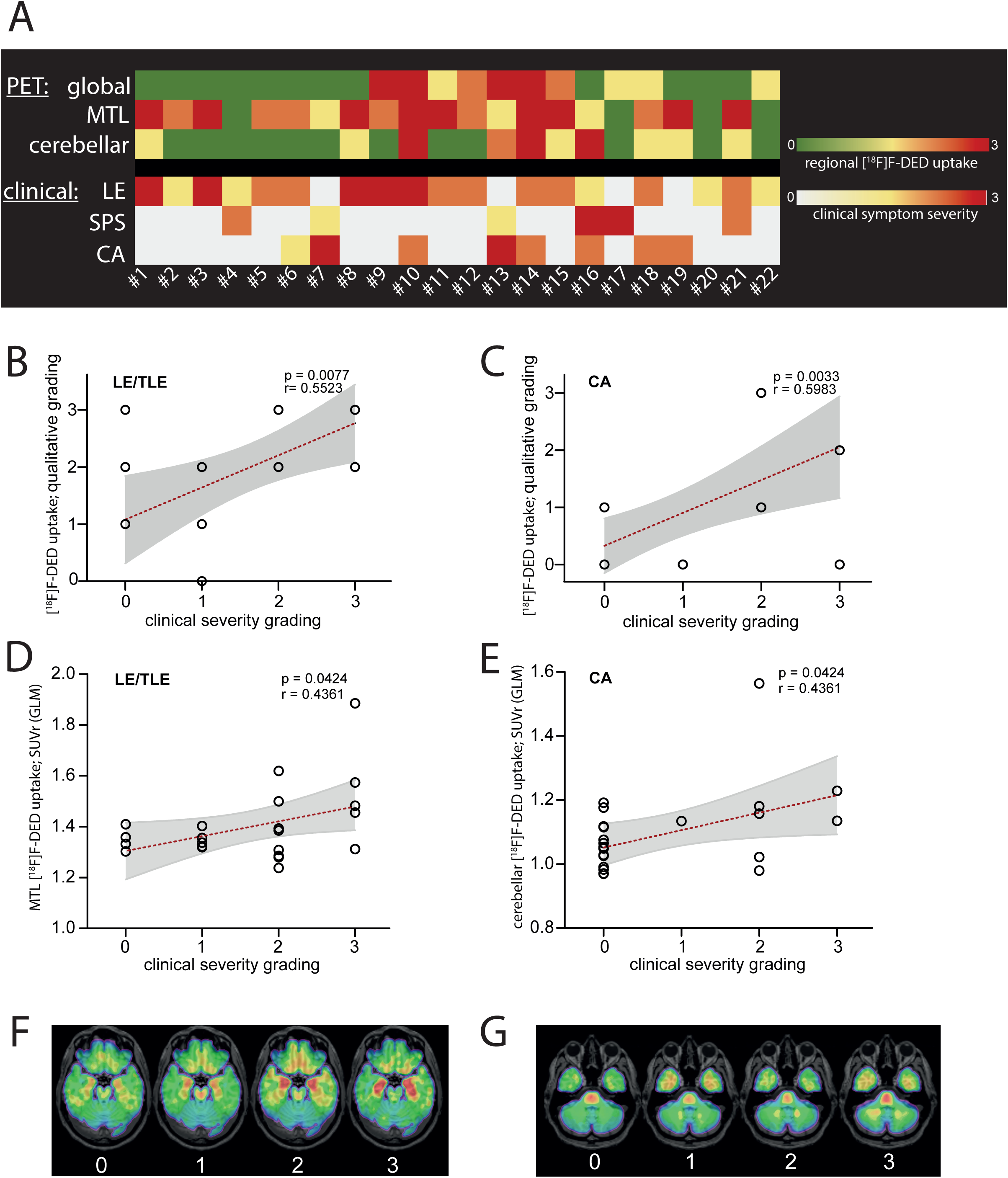
Correlation of regional [18F]F-DED uptake with clinical phenotype and severity. (**A**) Heatmap showing the relationship between clinical phenotype and severity with regional [^18^F]F-DED uptake in mesiotemporal (MT)/parahippocampal region, and cerebellar white matter. (**B**, **D**) Correlation of limbic encephalitis (LE)/ temporal lobe epilepsy (TLE) clinical severity with qualitative grading (**B**) and quantitative assessment of MT/parahippocampal [^18^F]F-DED uptake (standard uptake values [SUVr] with global mean scaling) (**D**) was assessed by linear regression. (**C**, **E**) Correlation of CA clinical severity with qualitative grading (**C**) and quantitative assessment of cerebellar white matter [^18^F]F-DED uptake ([SUVr] with global mean scaling) (**E**) was assessed by linear regression. (**F**) Representative patients showing different gradings of MT/parahippocampal tracer uptake: 0 - patient #7, 1 - patient #22, 2 - patient #6, 3 - patient #1. (**G**) Representative patients showing different gradings of cerebellar white matter tracer uptake: 0 - patient #22, 1 - patient #6, 2 - patient #18, 3 - patient #13.

We also assessed whether regional [^18^F]F-DED uptake correlate with clinical severity (grading from 0-3, as specified in the Methods section). We observed a significant correlation between clinical severity in LE and MT/ parahippocampal [^18^F]F-DED uptake, both through visual assessment of uptake (*p*=0.0077) (**Fig. 4B**) and quantitative analysis using SUVr GLM values (*p*=0.0424) (**Fig. 4D**). Similarly, clinical severity in CA (graded from 0-3, as specified in the Methods section) correlated with cerebellar white matter uptake, both qualitatively (*p*=0.0033) (**Fig. 4C**) and quantitatively (*p*=0.0424) (**Fig. 4E**). Representative [^18^F]F-DED PET images illustrating the different grading levels of MT/ parahippocampal as well as cerebellar white matter tracer uptake are presented in **Fig. 4F** and **4G**.

## Discussion

In this well-characterized cohort of 22 patients with GAD65-AIE, we evaluated the novel MAO-B PET radiotracer [^18^F]F-DED to asses reactive astrogliosis and characterize regional uptake patterns across distinct clinical phenotypes, with the aim of establishing a surrogate marker for disease activity. Our key findings are as follows:

- GAD65-AIE patients showed globally increased [^18^F]F-DED uptake compared to controls. Regional uptake was elevated in the MTL/parahippocampal region in LE/TLE patients, and in the cerebellum of CA patients.
- [^18^F]F-DED PET imaging demonstrated higher sensitivity for detecting MT abnormalities in GAD65-LE/TLE than conventional routine cMRI (83.3% [15/18] vs. 27.8% [5/18]).
- No significant correlation was identified between [^18^F]F-DED uptake and sNfL, but a correlation trend with sGFAP was noted.
- Regional [^18^F]F-DED signal intensity correlated with clinical severity in LE/TLE and CA phenotypes.

Emerging evidence supports a role for reactive astrogliosis in AIE. In a rat model, hippocampal astrocyte proliferation was induced following LGI1-IgG and NMDAR-IgG infusion ^15^. Histopathological studies of human hippocampal tissue have similarly revealed astrocytic activation in AIE ^16, 17,18^. A recent comprehensive neuropathological study – including AIE patients with Abs against both neuronal surface and intracellular antigens (including GAD65) – demonstrated increased densities of GFAP+ astrocytes and neurotoxic astrogliosis (GFAP⁺C3⁺) in the context of neuroinflammation ^19^. Prior immunohistochemistry studies in mice have shown a strong correlation between GFAP-positive astrocytes and MAO-B expression ^11^. Our findings extend these observations by demonstrating the in vivo detection of astrogliosis in GAD65-AIE using [^18^F]F-DED PET.

The observed regional correlation between [^18^F]F-DED uptake and clinical phenotypes, as well as areas of cMRI abnormalities ^20^, reinforces the specificity of the PET signal. Given its superior sensitivity compared to conventional MRI, [^18^F]F-DED may serve as a valuable adjunctive tool in diagnosing GAD65-AIE. Furthermore, it holds the potential for detecting subclinical or prodromal stages of overlapping phenotypes, thereby enabling earlier therapeutic intervention, as illustrated by the case (patient #21) presented in this study. Our findings also highlight the high prevalence of overlapping phenotypes in GAD65-AIE (9/22; 41%), supporting the concept of GAD65-associated neurological disorders as a disease spectrum ^3^.

Beyond aiding diagnosis and phenotype classification, [^18^F]F-DED PET is a promising tool for monitoring treatment response, as it reflects disease severity in LE/TLE and CA. GAD65-Ab titers, although diagnostically relevant, are not considered indicative of disease activity in GAD65-AIE ^3^. MRI findings are often unremarkable, and the disease course can be fluctuating. Thus, reliable paraclinical tools are critically needed.

Effective therapeutic strategies are especially needed for patients with GAD65-LE/TLE and GAD65-CA ^3^. Emerging treatment strategies include anti CD19-CART cells in SPS ^21^, anti-IL6 therapy (tocilizumab) in GAD65-LE ^22^, Janus kinase inhibitors ^23^, and T-cell depleting strategies ^24^. A robust imaging biomarker such as [^18^F]F-DED PET could enable precise monitoring of treatment efficacy. The proposed clinical composite scores to categorize clinical severity, adapted as a shortened and GAD65-AIE tailored version of the CASE score ^13^, could further improve clinical monitoring. In contrast, the modified Rankin Scale (mRS), so far commonly used to asses treatment outcomes, lacks the sensitivity to detect the relevant clinical findings in this condition ^25^.

Although we did not observe a strong correlation between regional [^18^F]F-DED uptake and serum biomarkers, our prior work showed elevated sNfL levels in early-stage GAD65-AIE, particularly in CA ^26^. The present study, conducted at a median disease duration of five years, may have missed such early sNfL elevations. Alternatively, [^18^F]F-DED uptake and sNfL increase may reflect different aspects of disease pathology. While increased sGFAP levels have been linked to disease progression in multiple sclerosis ^27^ and disease activity and disability in NMOSD ^28^, its role in AIE remains unclear. In this cross-sectional study, cerebellar [^18^F]F-DED uptake showed a trend toward association with sGFAP levels. Notably, voxel-wise analyses revealed an association between [^18^F]F-DED white matter signal and sGFAP levels. Larger studies are needed to confirm these preliminary findings. Together, [^18^F]F-DED PET along with measurement of sGFAP levels may form a useful composite for in vivo assessment of disease activity.

Limitations of the study include the relatively small sample size and the cross-sectional design, which may introduce confounding factors such as variable disease duration, episodes of disease reactivation, and ongoing immunosuppressive therapy. The use of heterogeneous MRI protocols precluded quantitative analysis of cMRI data. Nonetheless, our pilot findings support the feasibility and clinical relevance of [^18^F]F-DED PET in a well-defined cohort of GAD65-AIE patients. Future longitudinal studies with larger cohorts, standardized cMRI protocols, and longitudinal analyses, will help to validate these promising observations.

In conclusion, [^18^F]F-DED PET represents an encouraging imaging modality for in vivo assessment and monitoring of astrogliosis in GAD65-AIE. It holds potential as a biomarker for disease activity and a valuable tool to guide and assess therapeutic interventions.

### Study Funding

JA Gernert has received a research grant from the Deutsche Forschungsgemeinschaft (DFG, German Research Foundation; SFB/TRR 274, ID 408885537). G Höglinger and R Perneczky were funded by the Deutsche Forschungsgemeinschaft (DFG, German Research Foundation) under Germany’s Excellence Strategy within the framework of the Munich Cluster for Systems Neurology (EXC 2145 SyNergy – ID 390857198). M Brendel was funded by the Deutsche Forschungsgemeinschaft (DFG) under Germany’s Excellence Strategy within the framework of the Munich Cluster for Systems Neurology (EXC 2145 SyNergy—ID 390857198), the Alzheimer’s Drug Discovery Foundation, and the Michael J Fox Foundation (MJFF-021137). FS Thaler was funded by the Fritz Thyssen Stiftung (Az. 10.23.1.015MN), the Deutsche Forschungsgemeinschaft (DFG) under Germany’s Excellence Strategy within the framework of the Munich Cluster for Systems Neurology (EXC 2145 SyNergy—ID 390857198), and the Medical & Clinician Scientist Program (MCSP) of the LMU Munich.

### Disclosure

JS Dorneich reports no conflicts of interest; JA Gernert has participated in meetings sponsored by, received travel funding and non-financial support from Merck, Novartis, Roche, received honoraria for speaker engagements from WVAO eV and received a research grant from The Sumaira Foundation, all outside the present work; L Tagnin, M Zeinaki, L Sanzo, L Vogler, E Kaufmann, J Dargvainiene, and F Lexpoldt report no conflicts of interest; GN Bischof is an employee of Life Molecular Imaging; R Perneczky has received honoraria for advisory boards and speaker engagements from Roche, EISAI, Eli Lilly, Biogen, Janssen-Cilag, AstraZeneca, Schwabe, Grifols, Novo Nordisk, and Tabuk. He holds shares of Vistim Ltd. and Medotrax GmbH; B-S Rauchmann and S Lindner report no conflicts of interest; G Höglinger has ongoing research collaborations with Roche, UCB, Abbvie; serves as a consultant for Abbvie, Alzprotect, Amylyx, Aprinoia, Asceneuron, Bayer, Bial, Biogen, Biohaven, Epidarex, Ferrer, Kyowa Kirin, Lundbeck, Novartis, Retrotope, Roche, Sanofi, Servier, Takeda, Teva, UCB; received honoraria for scientific presentations from Abbvie, Bayer, Bial, Biogen, Bristol Myers Squibb, Esteve, Kyowa Kirin, Pfizer, Roche, Teva, UCB, Zambon; RA Werner received speaker honoraria from Novartis/AAA and PentixaPharm, and has performed advisory board work for Novartis/AAA and Bayer; M Kerschensteiner has been on advisory boards for Biogen, Merck, Novartis and Sanofi, has received speakers fees from Alexion, Biogen, Merck, Novartis, Sanofi, Roche and Teva and grant support from Sanofi and Roche (none in the context of this study); T Kümpfel has received personal fees for advisory boards from Alexion/Astra Zeneca, UCB, Merck and Biogen and for speaker honoraria/chairs and/or lectures/education from Alexion/Astra Zeneca, Novartis Pharma, Roche Pharma, Horizon Therapeutics/Amgen, Chugai Pharma. The Institution she works for has received compensation for serving as a member of a steering committee from Roche. She is a site principal investigator in several randomized clinical trials (Novartis Pharma, Roche Pharma, BMS and Sanofi Genzyme) and in a randomized clinical trials supported by the BMBf (funding code: 01GM1908E) and her institution has received compensation for clinical trials all outside the present work; M Brendel received speaker honoraria from GE healthcare, Roche, and Life Molecular Imaging and advised GE healthcare, MIAC, and Life Molecular Imaging; FS Thaler received grant support from Novartis Pharma GmbH and speaker honoraria from Alexion.

## Supporting information

eFigure 1

## Data Availability

Data are available to qualified researchers upon reasonable request.

## Abbreviations

Abs: antibodies
AIE: autoimmune encephalitis
CA: cerebellar ataxia
[^18^F]F-DED: [^18^F]fluorodeprenyl-D2
GAD65: glutamic acid decarboxylase 65
sGFAP: serum glial fibrillary acidic protein
LE: limbic encephalitis
MAO-B: monoamine oxidase B
MT: mesiotemporal
PET: positron-emission-tomography
sNfL: serum neurofilament light chain
SUV: Standardized uptake values
VT: volumes of distribution
SPS: stiff person syndrome
TLE: temporal lobe epilepsy

## Acknowledgments

Precursor of [^18^F]F-DED was kindly provided by Life Molecular Imaging

## Figure legends

**eFigure 1**: *Methodological comparison of [^18^F]F-DED uptake quantification.* (A) Quantification was performed using static acquisition (30-60 min post-injection) and kinetic modelling of the full 60 min imaging window. Standardized uptake values (SUV) were determined and normalized to global cortex and subcortex values (SUVr GLM). Averaged SUVr GLM image data of all GAD65-AIE patients (n=22) and controls (n=8) are shown. (B) Correlations between global mean scaled kinetic modelling (VT, 1TC2k model) and global mean scaled SUVr (GLM) in cortex, cerebellar white matter, and MT/ parahippocampal region are shown and were assessed by linear regression.

**eFigure 2:** *Evaluation of effect of immunotherapy and disease duration on [^18^F]F-DED uptake*. **(A)** [^18^F]F-DED uptake (standard uptake values [SUVr] with global mean scaling) in the cortex, mesiotemporal (MT)/parahippocampal region, and cerebellar white matter was compared between patients with immunotherapy and without immunotherapy, and an unpaired T-Test with Welch-correction was applied. (**B**, **C**) Correlation of [^18^F]F-DED uptake (SUVr with global mean scaling) in the MT/parahippocampal region (**B**) and cerebellar white matter (**C**) with disease duration at the time of PET imaging was assessed by linear regression.

