## Supplementary figures and images for "[^18^F]fluorodeprenyl-D2 PET imaging as a novel tool to monitor disease activity in GAD65-Ab Autoimmune Encephalitis"

### e Figure 2

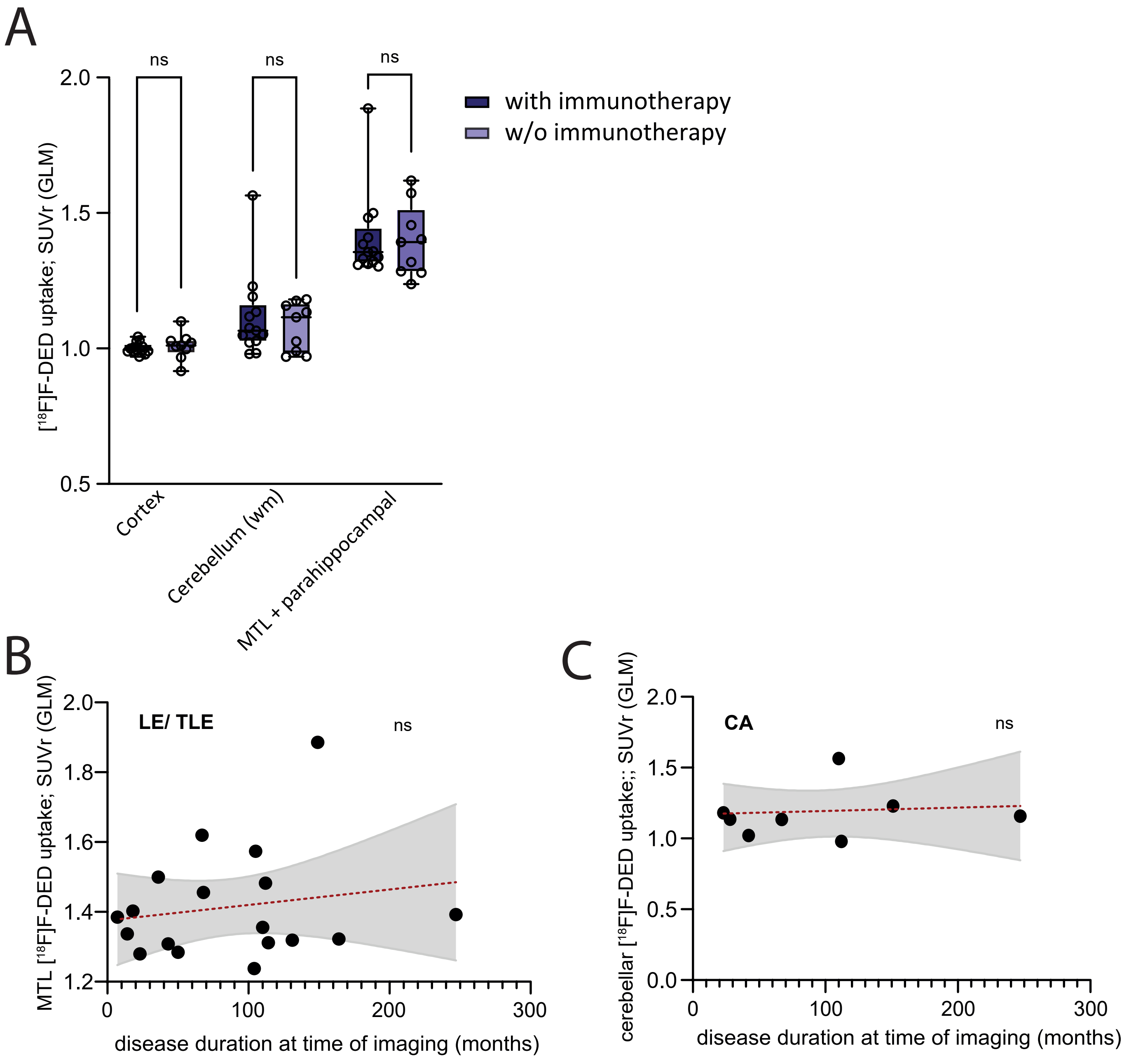

### eFigure 1

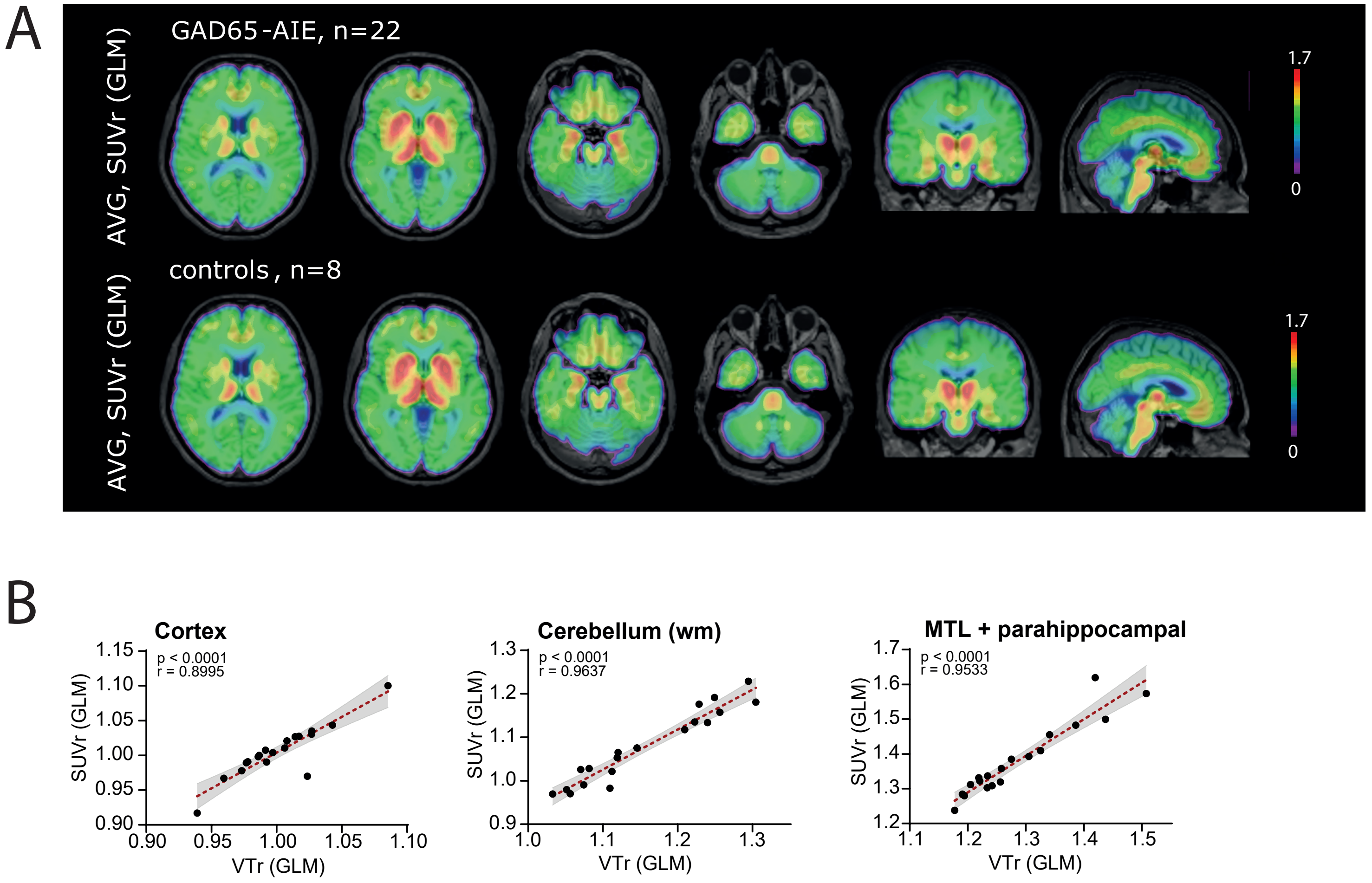
